# Identification of Ultra-Rare Variants Associated with Dilated Cardiomyopathy: Insights into LMNA Pathogenesis and Prognostic Implications including E115M and R190W

**DOI:** 10.1101/2025.08.01.25332667

**Authors:** Takeshi Kitai, Koichi Ishida, Yoshihiko Ikeda, Takuya Watanabe, Takuma Sato, Kanako Teramoto, Shinichi Kurashima, Shin Ito, Akihiro Asakura, Yuki Kuramoto, Shigemiki Omiya, Yasumasa Tsukamoto, Chisato Izumi, Kinta Hatakeyama, Tomohiro Nishinaka, Masao Nagasaki, Yasuhiko Sakata

## Abstract

**Background:** Although the genetic backgrounds of dilated cardiomyopathy (DCM) have been intensively investigated, the clinical impact of ultra-rare genetic variants (URVs) on DCM remains underexplored.

**Aims:** This study aimed to identify genes enriched for pathogenic coding URVs in DCM patients and elucidate their clinical and structural significance, with focus on *LMNA* mutations.

**Methods:** Among patients with DCM, the significance of pathogenic URVs (minor allele frequency ≤0.001%), identified based on predictions by AlphaMissense or protein truncation, was examined.

**Results:** Among 245 Japanese patients with DCM (53.1 ± 15.1 y.o., 18.0% female.), 43 patients (17.6%) had a total of 49 pathogenic URVs; 14 missense URVs in *LMNA* (10 patients), 3 missense URVs in *SLC51A* (3 patients), and 32 protein-truncating URVs in *TTN* (32 patients). Among the patients with URVs in *SLC51A*, one also carried URVs in *LMNA*, and another in *TTN*. Compared to those without any URVs, the *LMNA* URV carriers had increased risk of the composite of cardiovascular death, heart transplantation, and left ventricular assist device implantation (hazard ratio [HR] 6.44, 95% CI: 2.85-14.54; P<0.001) as well as implantable cardioverter defibrillator implantation (HR 4.49, 95%CI: 2.02-9.94; P<0.001). Among them, seven amino acid substitutions were identified, including E115M and R190W, which were found in three patients each. Six of the substitutions were localized to the 1B and 2B domains of the alpha-helical coiled-coil structure that play a role in modulating the elasticity of lamin. Histological examinations revealed abnormal nuclear morphology in cardiomyocytes of *LMNA* URV carriers.

**Conclusion:** Pathogenic URVs were identified in 17.6% of patients with DCM. Among them, the *LMNA* URV carriers were characterized by histologically abnormal nuclear structure in cardiomyocytes and increased risk of adverse cardiovascular events.

**CLINICAL PERSPECTIVE:** *What’s New?:*

- This is the first comprehensive investigation to delineate both the clinical and histopathological significance of coding ultra-rare variants (URVs; allele frequency ≤0.001% in control populations) in dilated cardiomyopathy (DCM).
- Utilizing whole-genome sequencing in a Japanese cohort of 245 patients with DCM, this study identified pathogenic coding URVs in 17.6% of cases, with a predominance in *LMNA* and *TTN*.
- Carriers of *LMNA* URVs—including those with a potentially population-enriched variant (E115M)—demonstrated a significantly elevated risk of severe cardiovascular events and exhibited distinctive nuclear abnormalities in cardiomyocytes.

*What are the clinical implications?:*

- Genetic screening for *LMNA* URVs in patients with DCM may enable early identification of individuals at elevated risk for adverse cardiovascular outcomes.
- These findings support the need for genotype-guided clinical management, including early initiation of guideline-directed medical therapy and consideration of prophylactic implantable cardioverter-defibrillator (ICD) placement in *LMNA* URV carriers.
- Identification of population-specific variants, such as E115M, may enhance risk stratification and contribute to improved long-term prognosis through tailored surveillance and intervention strategies.

## INTRODUCTION

As the global population continues to age, the prevalence of heart failure (HF) has risen sharply, highlighting the need for innovative approaches to its diagnosis and management.^1^ Dilated cardiomyopathy (DCM) is a leading cause of HF, affecting a substantial proportion of patients and characterized by impaired myocardial contractility and left ventricular enlargement.^2–4^ Conventional diagnostic approaches, which rely primarily on imaging, histology, and clinical profiling, often fail to elucidate underlying etiologies or guide cause-specific therapeutic strategies. As a result, despite recent advances in pharmacologic and device-based therapies, the quality of medical care, particularly in the context of personalized medicine, remains suboptimal among patients with DCM.

Advances in genomic technologies, particularly whole-genome sequencing (WGS), have significantly deepened our understanding of the genetic underpinnings of cardiomyopathies, including DCM, and have paved the way for the development of personalized medical strategies.^5,6^ On the other hand, genetic research has predominantly focused on common variants with well-established links to diseases.^7,8^ Furthermore, polygenic risk scores based on common variants have successfully identified individuals at high risk for certain diseases. However, emerging studies have highlighted the importance of ultra-rare variants (URVs), which provide clearer genetic causality compared to common variants.^9,10^

Due to their rarity, URVs can pinpoint specific genetic abnormalities that contribute to disease pathogenesis, offering more precise insights into the genetic basis of diseases.^11–14^ The recent release of large-scale sequencing data from the UK Biobank has substantially expanded the scope of URV detection and analysis. Leveraging exome sequencing data from 269,171 individuals of European ancestry, they systematically investigated the association between rare variants and a comprehensive set of phenotypes. As a result, 1,703 genes were identified as being associated with phenotypes. These associations were significantly enriched for targets of therapeutics approved by the U.S. Food and Drug Administration, strongly suggesting that investigating URVs is highly valuable for elucidating disease mechanisms and identifying novel therapeutic targets.^15^ This precision by URVs has the potential to provide a more detailed understanding of the genetic architecture of cardiomyopathy, enabling the development of tailored therapeutic interventions. Nevertheless, despite these advancements, the role of URVs in DCM remains largely unexplored, particularly in terms of their clinical and mechanistic significance.

In this study, we aimed to investigate the frequency and clinical implications of URVs in patients with DCM, using data from the National Cerebral and Cardiovascular Center (NCVC) Biobank (Suita, Japan) and the Tohoku Medical Megabank (TMM) dataset (Sendai, Japan).^16^ The 54K-individual general population WGS dataset from TMM (TMM54K) served as the control dataset, comprising WGS data from 54,302 unrelated Japanese individuals (108,604 alleles). These individuals were selected from a larger cohort of 69,014 participants enrolled in the TMM Project, a population-based initiative designed to advance personalized medicine through comprehensive genomic and multi-omics analysis. To minimize bias in allele frequency estimation, related individuals were systematically excluded. The cohort spanned a broad age range and included both sexes, representing the general Japanese population. By leveraging AlphaMissense predictions and protein-truncating variant analysis, we aimed to advance our understanding of URVs and their potential role in the personalized management of DCM.

## METHODS

### Study design and patient population

Patients with DCM who participated in the NCVC Biobank study between 2012 and 2020 and underwent WGS in 2020 were retrospectively analyzed. The diagnosis of DCM was based on the clinical judgment of specialists for HF at the NCVC and was comprehensively determined for patients with non-ischemic HF using echocardiography, cardiac magnetic resonance imaging (MRI), and endomyocardial biopsy. In this study, we set the date of blood collection as the reference point (Day 0) and the date of birth as an anchor for age calculation. On Day 0, the ages of the subjects ranged from approximately 19 to 87 years. The primary endpoint was the composite consisting of cardiovascular (CV) death, heart transplantation (HTx), or left ventricular assist device (LVAD) implantation. The secondary endpoints consisted of all-cause death, CV death, HTx, LVAD implantation, HF hospitalization, and implantation of a cardioverter defibrillator, namely, either an implantable cardioverter defibrillator (ICD) or a cardiac resynchronization therapy defibrillator (CRT-D). The outcome data were collected using electronic medical records. Clinical characteristics and outcomes were compared by the presence or absence of the URVs identified.

The study was conducted in compliance with the Declaration of Helsinki and Japanese Ethical Guidelines for Medical and Health Research involving Human Subjects. The study protocols of the NCVC Biobank study and the present study were approved by the ethics committee of the NCVC (Research Approval Number: M23-068 and R21059, respectively). All patients provided written informed consent to the NCVC Biobank study at the time of registration, and the information disclosure on the NCVC website guaranteed their opportunities for refusal to participate in the NCVC Biobank study and the present study.

### Whole genome sequencing

DNA samples from 245 DCM patients were prepared immediately after blood collection for the NCVC Biobank study using manufacturer-recommended procedures and reagents.^17^ Sequence libraries were performed according to the protocol of The TruSeq DNA PCR-Free HT Library Prep Kit (Illumina Inc., California, USA). Sequencing was performed on an Illumina NovaSeq 6000 platform (Illumina Inc., California, USA) to generate 150-bp paired-end reads, multiplexed to yield > 90Gb per sample after duplicate removal.

In this study, variant analysis was followed the workflow described in Table 2 and Supplementary Table 1 of our prior publication.^18^ Briefly, high-quality reads filtered from FASTQ files were mapped to the human reference genome GRCh38DH using BWA (v0.7.17). Variant calling was conducted using GATK (v4.1.4) HaplotypeCaller, and joint-genotyping was performed with GenomicDBImport and GenotypeGVCFs across 3,169 genomic intervals. For the generated population gVCF (variant call format) file, VQSR operation is applied to set the quality scores to variants to produce the final VCF. For the VCF, annotation was applied with a tool SnpEff (v4.3i). This gave genomic location, type of mutation, functional impact, and other associated genetic information for the following downstream analysis.

### Analysis of URVs

The procedures to select coding pathogenic candidate URVs accumulating gene in 245 DCM samples were illustrated in **Supplemental Figure 1**. We utilized custom scripts to analyze the WGS data, identifying pathogenic candidate URVs with minor allele frequencies (MAF) of ≤0.001% in TMM54K.^16^

The pathogenic candidate URVs were selected based on two different protein functional effect or protein expression criteria: (i) likely-pathogenic missense variants (scores ≥0.564) predicted by AlphaMissense (DeepMind Technologies Ltd., London, UK)^19,20^ or (ii) protein-truncating variants annotated by SnpEff.

To perform gene-based enrichment analysis, Fisher’s exact test was conducted to compare allele counts of candidate URVs from criteria (i) or (ii) between DCM cases and the 54KTMM controls, and odds ratios were calculated. For genes demonstrating significant enrichment, clinical outcomes analyses were performed in patient subgroups carrying the corresponding candidate URVs.

### Three-dimensional structure of *LMNA* mutations

Amino acid substitutions resulting from the pathogenic candidate URVs in *LMNA* were mapped onto Alphafold2-generated structural models (DeepMind Technologies Ltd., London, UK)^21^, and their precise locations within defined *LMNA* domains were annotated.

### Histological analysis in myocardial specimens from patients with and without *LMNA* URVs

For histological comparison of nuclear structural morphology in myocardial specimens, ten control cases without any pathogenic *LMNA* variant, either ultra-rare or common, were selected after being matched for age and sex with *LMNA* URV patients at the time of blood collection. Images of myocardial specimens from the right or left ventricle via endomyocardial biopsy during diagnostic evaluation or LVAD implantation were available in nine out of ten patients with *LMNA* URVs and eight of ten control cases.

### Statistical analysis

Continuous measures were indicated as mean ± standard deviation or median (interquartile range). Categorical measures were indicated as n (%). P values are based on Kruskal-Wallis test, χ-square test or Fisher’s exact test. Two-sided P values <0.05 were considered statistically significant. While URVs were defined using a MAF cutoff of ≤0.001% in the primary analysis, a sensitivity analysis was additionally conducted using a less stringent threshold of ≤0.01%.

Cumulative incidence of outcomes was estimated in patient subgroups carrying the corresponding candidate URVs, and the differences between groups were evaluated using Gray’s test with all-cause death as a competing risk. Cumulative incident curves were additionally generated for visualizing overall survival. The univariable and multivariable Cox regression analyses were performed to evaluate the association between variables and outcomes.

Fisher’s exact test was used for the pathogenic URV enrichment gene-based comparison between DCM cases and TMM54K controls. Genes with Bonferroni-corrected significance (P<0.05/19,872; equivalent to −log_10_(p) >5.60) were considered significant. Statistical analyses were conducted in R (v4.2.1).

## RESULTS

### Patient characteristics

A total of 245 patients with DCM were analyzed in this study. Clinical characteristics are summarized in **Table 1**. Among the 245 DCM patients, 201 (82.0%) were male, with a mean age of 53.1 ± 15.1 years at Day 0. **Table 2** showed the incidence of the primary and secondary endpoints. During the study through February 2024, 68 patients (27.8%) met the primary endpoint. Additionally, 29 patients (11.8%) died from all-causes, 199 patients (81.2%) were hospitalized for HF, and 101 patients (41.2%) received ICD/CRT-D implantation.

**Table 1.**
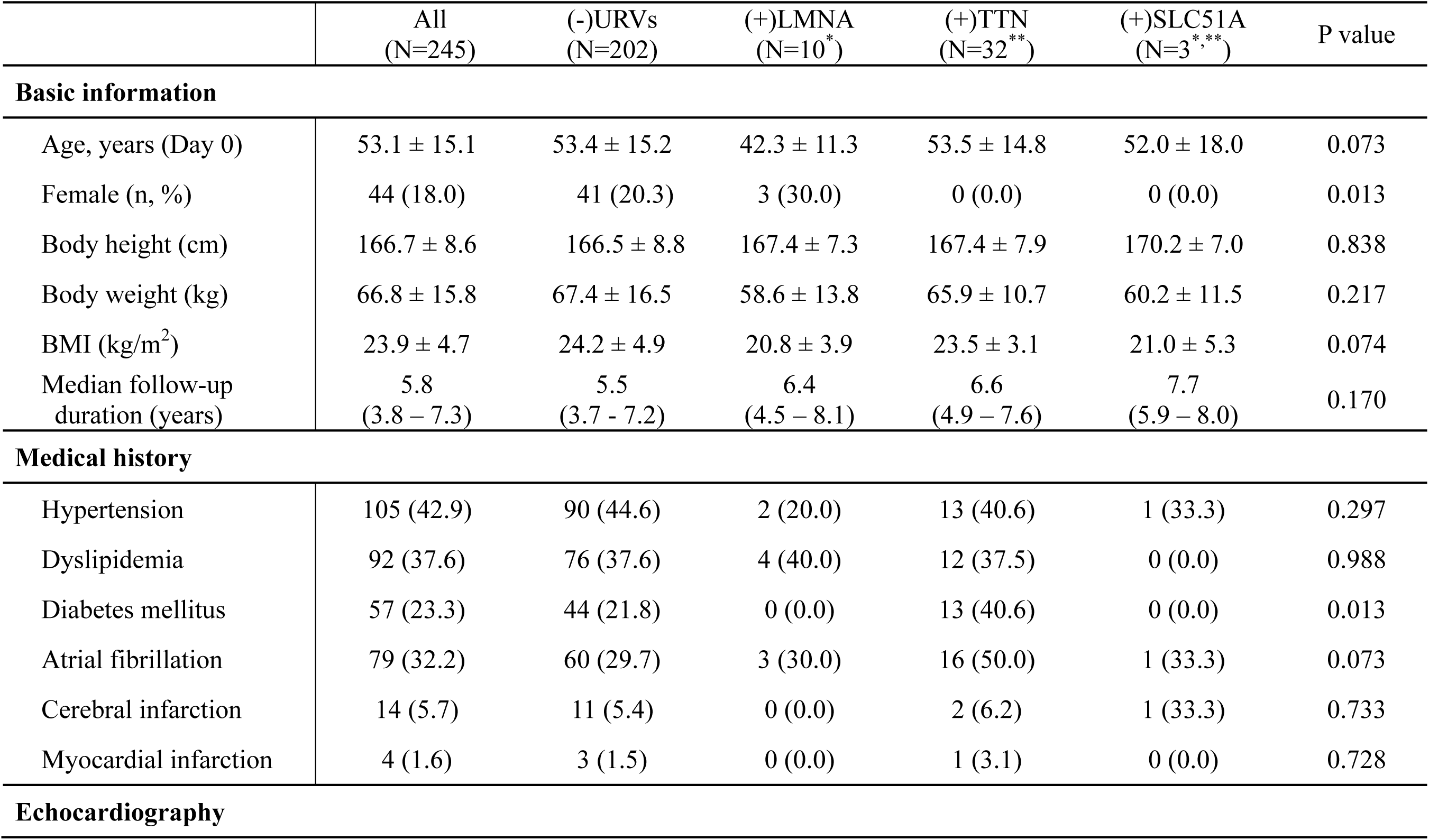
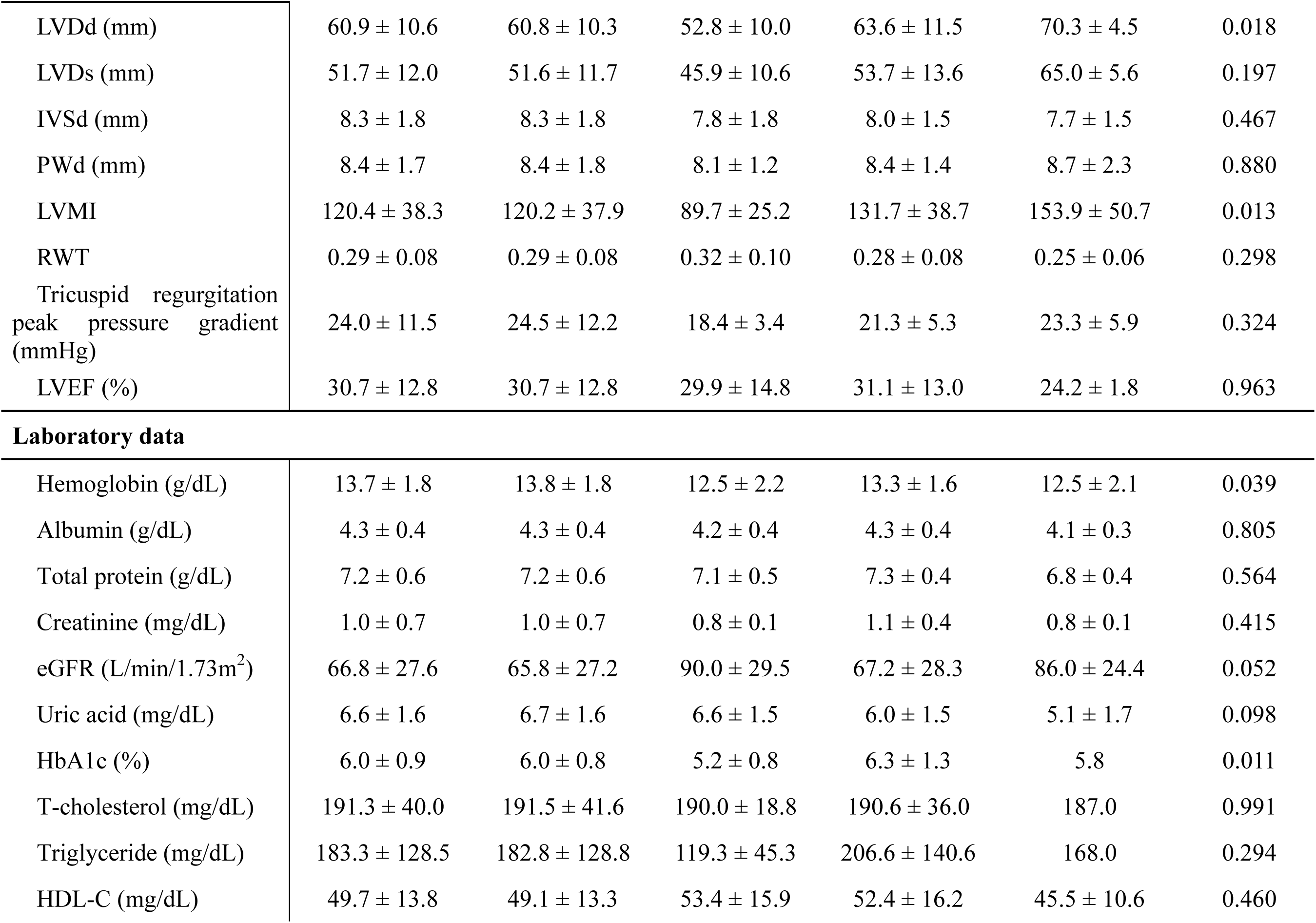
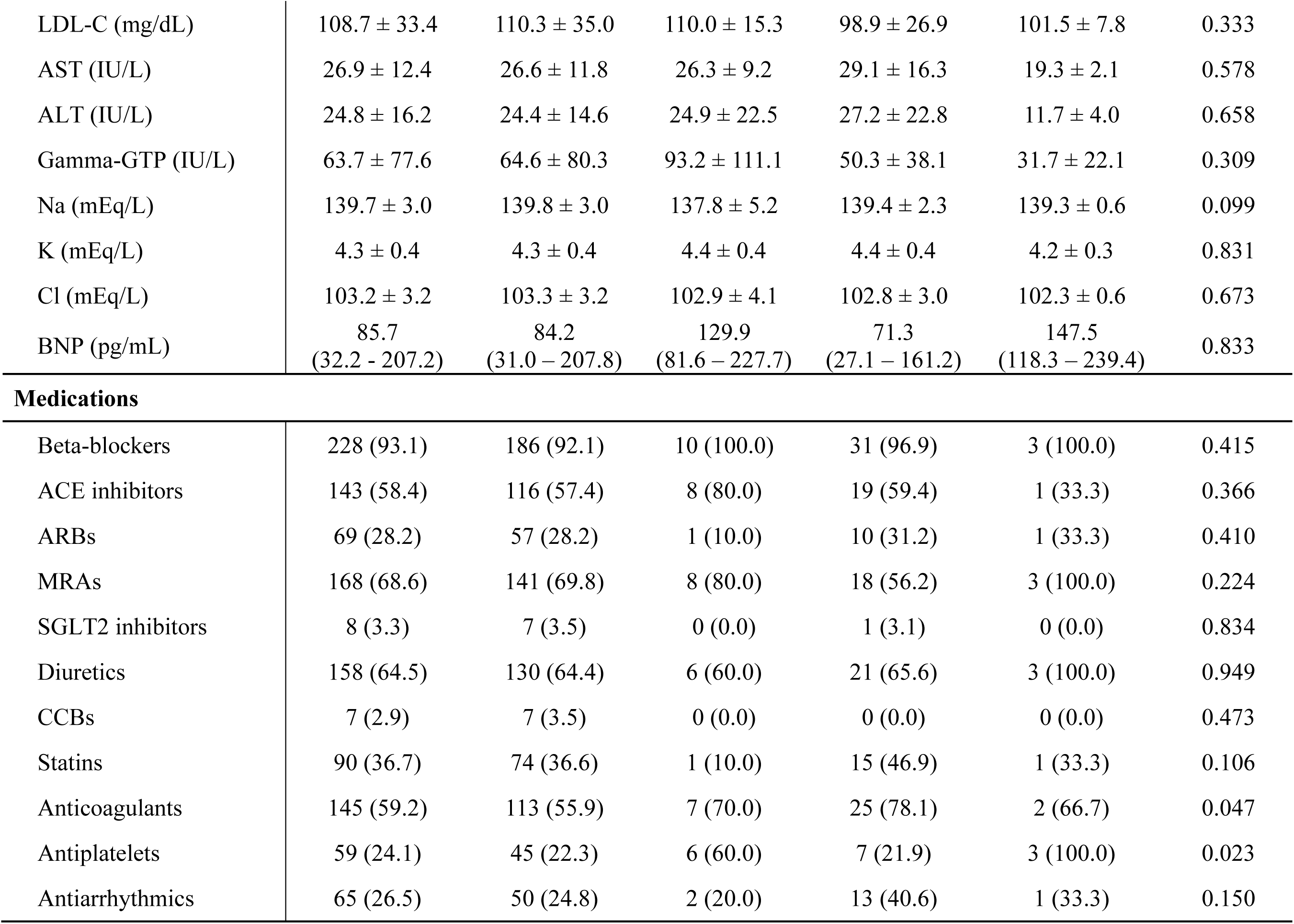
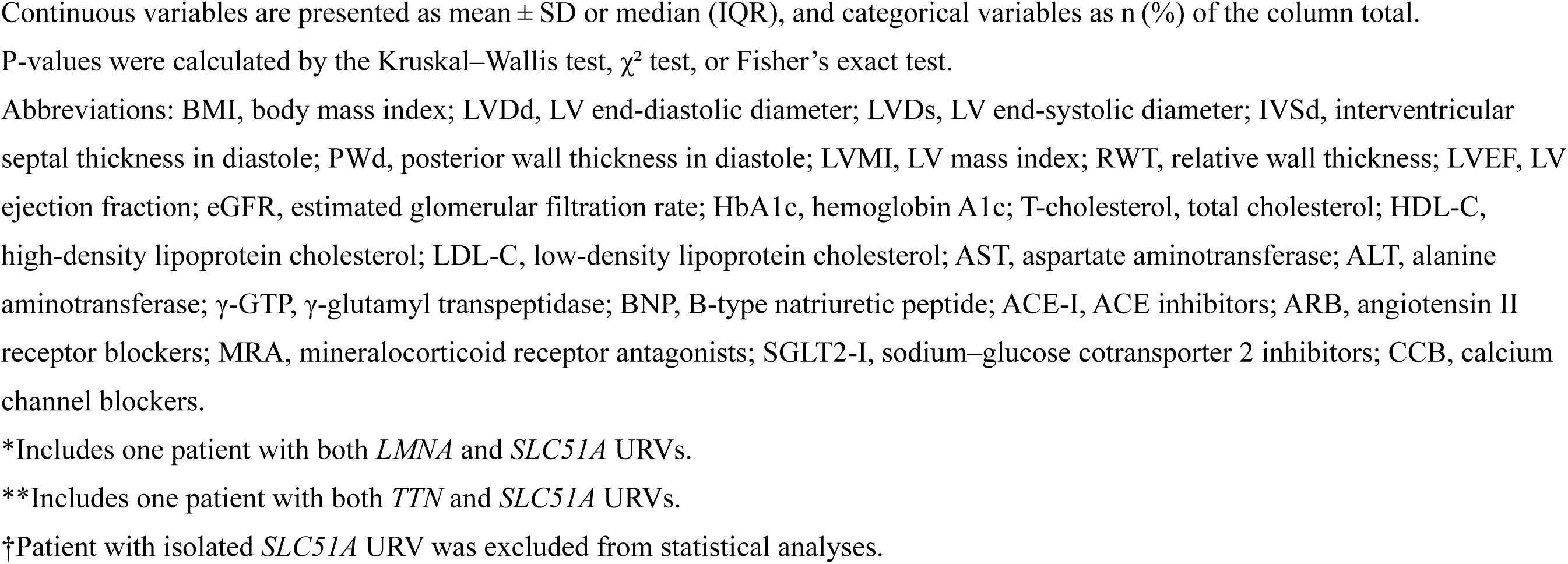
Baseline characteristics.

**Table 2.** Clinical outcomes of the study subjects.

|  | All<br>(N=245) | (-)URVs<br>(N=202) | (+)LMNA<br>(N=10) | (+)TTN<br>(N=32) | P value |
| --- | --- | --- | --- | --- | --- |
| Primary endpoint | 68 (27.8) | 49 (24.3) | 7 (70.0) | 12 (37.5) | 0.003 |
| - CV death | 20 (8.2) | 15 (7.4) | 0 (0.0) | 5 (15.6) | 0.183 |
| - HTx | 24 (9.8) | 17 (8.4) | 4 (40.0) | 3 (9.4) | 0.005 |
| - LVAD implantation | 49 (20.0) | 34 (16.8) | 7 (70.0) | 8 (25.0) | <0.001 |
| All-cause death | 29 (11.8) | 22 (10.9) | 0 (0.0) | 7 (21.9) | 0.101 |
| HF hospitalization | 199 (81.2) | 159 (78.7) | 9 (90.0) | 30 (93.8) | 0.099 |
| ICD or CRT-D implantation | 101 (41.2) | 80 (39.6) | 7 (70.0) | 13 (40.6) | 0.162 |
| Pacemaker implantation | 16 (6.5) | 14 (6.9) | 1 (10.0) | 0 (0.0) | 0.277 |

Categorical variables are presented as n (%). P-values were calculated by the $\chi^2$ test or Fisher's exact test.
Primary endpoint was a composite of cardiovascular death (CV death), heart transplantation (HTx), or left ventricular assist device (LVAD) implantation.
Abbreviations: CV, cardiovascular; HTx, heart transplantation; LVAD, left ventricular assist device; ICD, implantable cardioverter-defibrillator; CRT-D, cardiac resynchronization therapy defibrillator.

### Identification of pathogenic URVs to DCM with gene-based enrichment analysis

After QC filtering against the GRCh38 reference, 71,297 missense and 3,904 protein-truncating variants were detected across 245 DCM genomes. Of these, 7,260 likely pathogenic missense variants met likely-pathogenic criteria, and applying an ultra-rare threshold (MAF ≤0.001% in the TMM54K controls) refined the set to 351 missense URVs and 250 truncating URVs. Gene-based enrichment analysis (Fisher’s exact test with Bonferroni correction, –log_10_ p-value > 5.60) comparing DCM cases to TMM54K controls identified three enriched genes: *LMNA*, *SLC51A*, and *TTN*. In total, 43 patients (17.6%) harbored 49 pathogenic URVs: 14 *LMNA* missense URVs in 10 patients, three *SLC51A* missense URVs in three patients, and 32 *TTN* truncating URVs in 32 patients (**Table 3**).

**Table 3.** Genes Enriched in DCM Based on URVs with MAF ≤0.001%.

| Gene name | Amino acids | DCM cases |  | 54KTMM controls |  | p-value (-log10) | Odds ratio (95%CI) |
| --- | --- | --- | --- | --- | --- | --- | --- |
|  |  | Pathogenic allele count | Total allele count | Pathogenic allele count | Total allele count |  |  |
| Pathogenic missense mutations |  |  |  |  |  |  |  |
| LMNA | 664 | 14 | 490 | 20 | 108604 | 23.84 | 159.68<br>(80.18 – 318.01) |
| SLC51A | 340 | 3 | 490 | 1 | 108604 | 6.45 | 669.01<br>(69.47 – 6443.10) |
| Protein-truncating |  |  |  |  |  |  |  |
| TTN | 34350 | 32 | 490 | 163 | 108604 | 39.17 | 46.48<br>(31.47 – 68.65) |

Of the 43 patients, one had two URVs in *LMNA*, while one had a URV in both *LMNA* and *SLC51A*, and another had a URV in *SLC51A* and *TTN*. Because the effects of *SLC51A* URVs could not be separated from those of coexisting *LMNA* or *TTN* URVs, and because the sole patient with an isolated *SLC51A* URV did not meet the primary endpoint (**Supplemental Table 1**), this patient was excluded from further analyses. Subsequent analyses thus focused on the independent contributions of *LMNA* and *TTN* variants to patient outcomes. Since no patients had both *LMNA* and *TTN* URVs, we categorized 244 patients into three groups: (+)LMNA for those with *LMNA* URVs (N=10), (+)TTN for those with *TTN* URVs (N=32), and (-)URVs for those without any URV (N=202) (**Table 1**).

### Genotype-Phenotype Associations in DCM Patients

Among (+)LMNA patients, 70% (7/10) were male. The (+)LMNA patients were characterized by younger age, lower BMI, smaller left ventricular size, less frequent left ventricular hypertrophy, and higher usage of antiplatelet therapy, compared to (-)URVs. All (+)TTN patients were male (32/32) (fisher-exact test; P=0.004) and had a higher prevalence of diabetes mellitus and usage of anticoagulant therapy.

Among the three groups, (+)LMNA had a higher incidence of the primary and secondary endpoints (**Figure 1**, **Table 4**). The cumulative incidence curves for the primary endpoint and ICD/CRT-D implantation demonstrated a marked divergence among the three groups over time. The cumulative incidence of the primary endpoint and ICD/CRT-D implantation from birth was higher in (+)LMNA than (-)URVs (Gray’s test; P<0.001 and P=0.001, respectively). The Cox regression analyses showed that (+)LMNA had a higher risk of the primary endpoint (hazard ratio [HR] 6.44, 95% confidence interval [CI] 2.85-14.54; P<0.001), HF hospitalization (HR 2.28, 95% CI 1.15-4.48; P=0.018) and ICD/CRT-D implantation (HR 4.49, 95% CI 2.02-9.94; P<0.001), as compared to (-)URVs. After adjustment for year of birth, sex, and BMI, (+)LMNA still had a higher risk of the primary endpoint (HR 2.87, 95% CI 1.24-6.63; P=0.013) and LVAD implantation (HR 2.92, 95% CI 1.24-6.89; P=0.014) compared to (-)URVs. In contrast, (+)TTN was not associated with the risk of the primary endpoint compared to (-)URVs.

**Figure 1.**
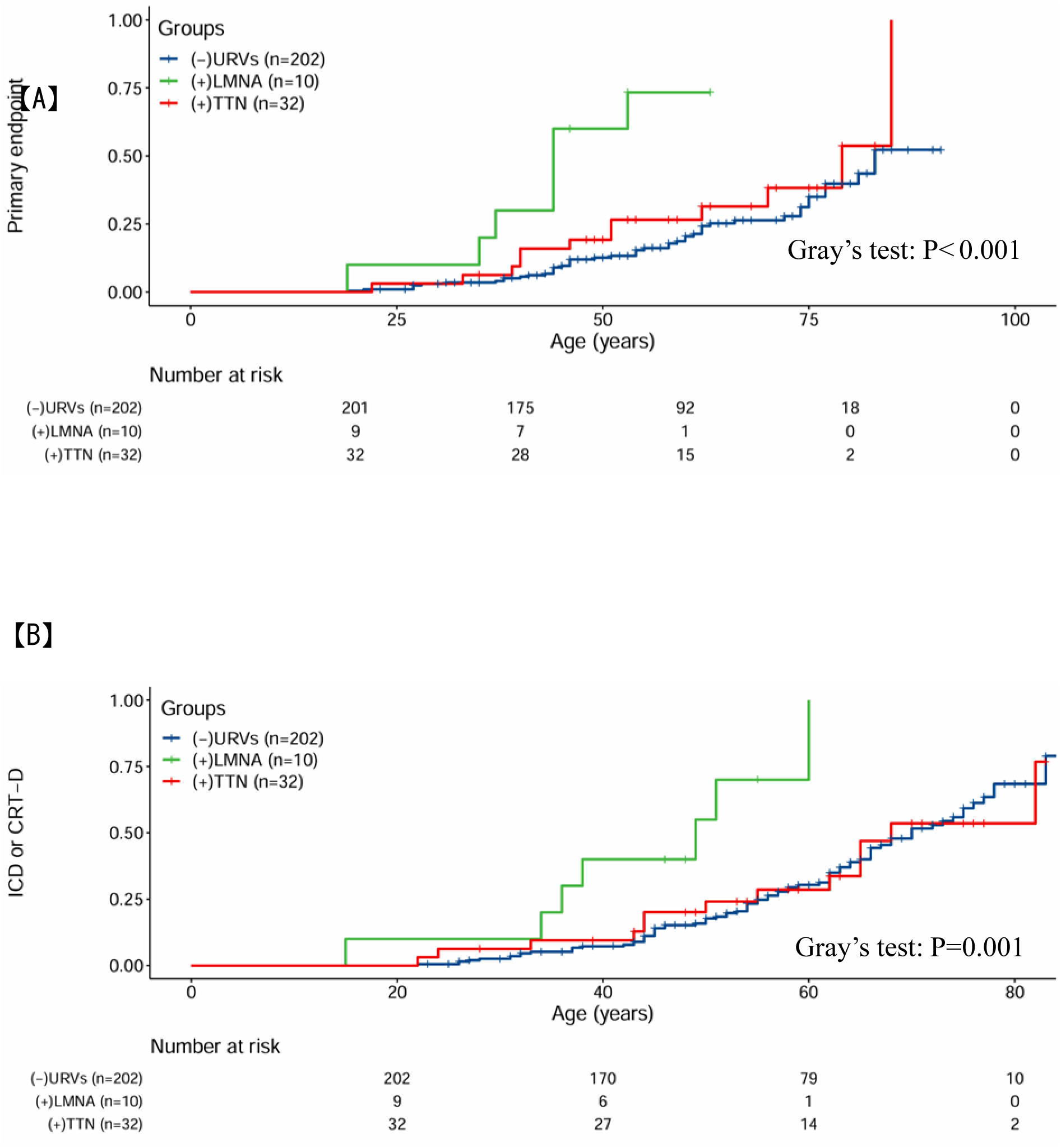
Cumulative incidence of clinical endpoints and device therapy. Cumulative lifetime incidence curves of (A, B) the primary endpoint (cardiovascular death, heart transplantation, or LVAD implantation) and (C, D) appropriate device therapy (ICD or CRT-D) in DCM patients with or without URVs. The x-axis shows age (years after birth), and the y-axis shows cumulative incidence. Statistical comparisons were made by Gray’s test; p-values are indicated in the panels.

**Table 4.**
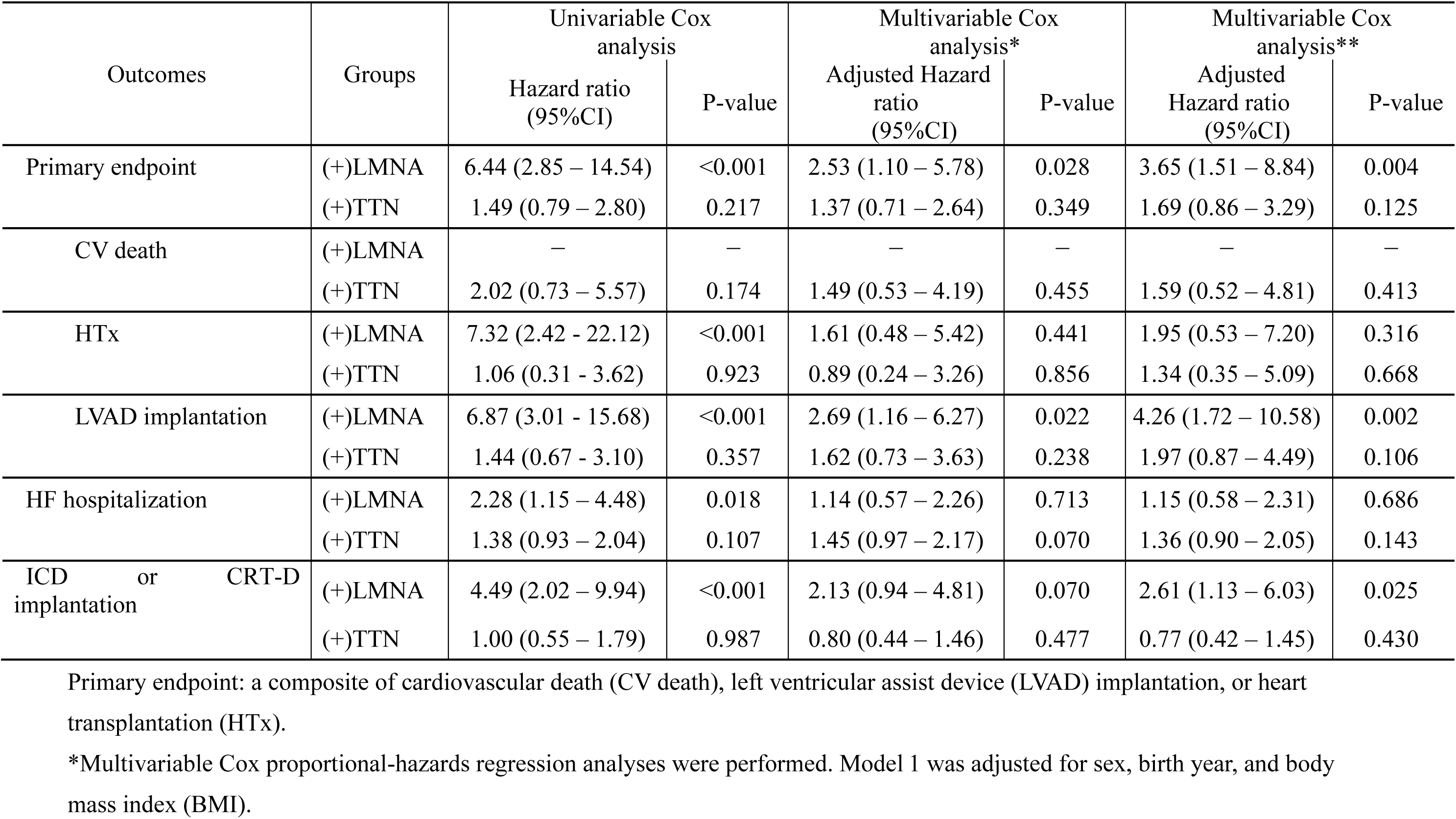
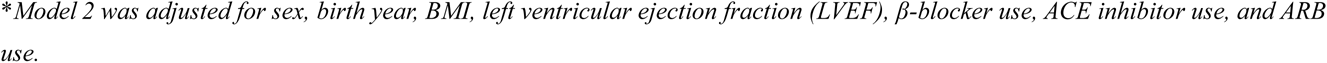
Cox regression analysis of ultra-rare variants in *LMNA* and *TTN* on Clinical Outcomes.

### Locations of the URVs in *LMNA* and Nuclear structural abnormalities

Among ten patients with (+)LMNA, eight unique URVs were identified in *LMNA* (**Table 5**). Among them, three patients had two consecutive URVs at positions 156115261 and 156115262 in Chromosome 1, causing an E115M variant, while another three patients had a URV at position 156134457 in Chromosome 1, causing an R190W variant. One patient had two different URVs, causing variants of L197R and T199I, respectively, and the remaining three patients had different URVs, causing variants of R335W, L363P, and R541H, respectively. Among the seven amino acid substitutions caused by the eight URVs, four and two were located in the Coil 1B and 2B regions of the *LMNA* gene, respectively (**Figure 2**).

**Table 5.**
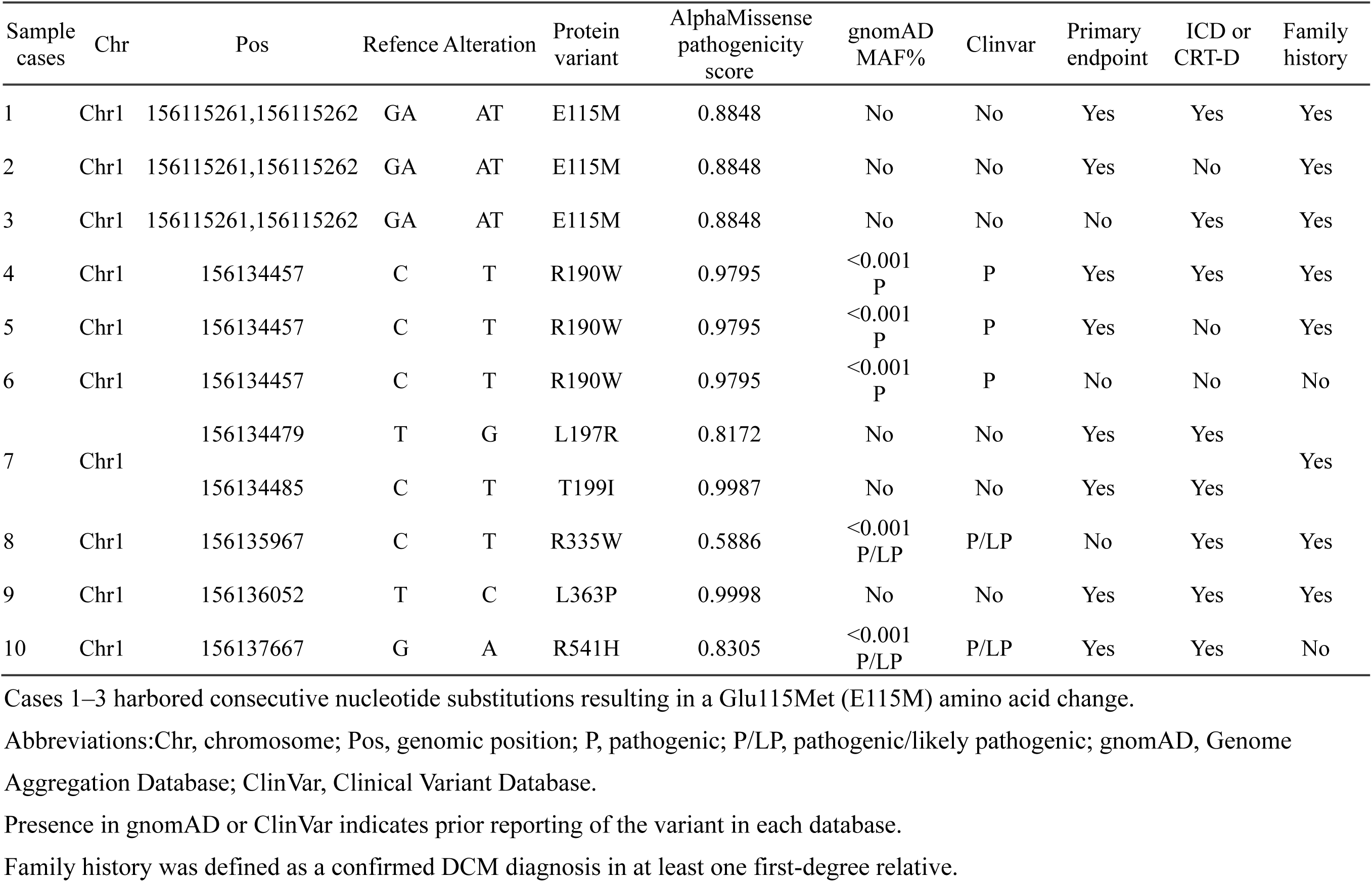
Characteristics of *LMNA* URVs identified in ten DCM patients.

**Figure 2.**
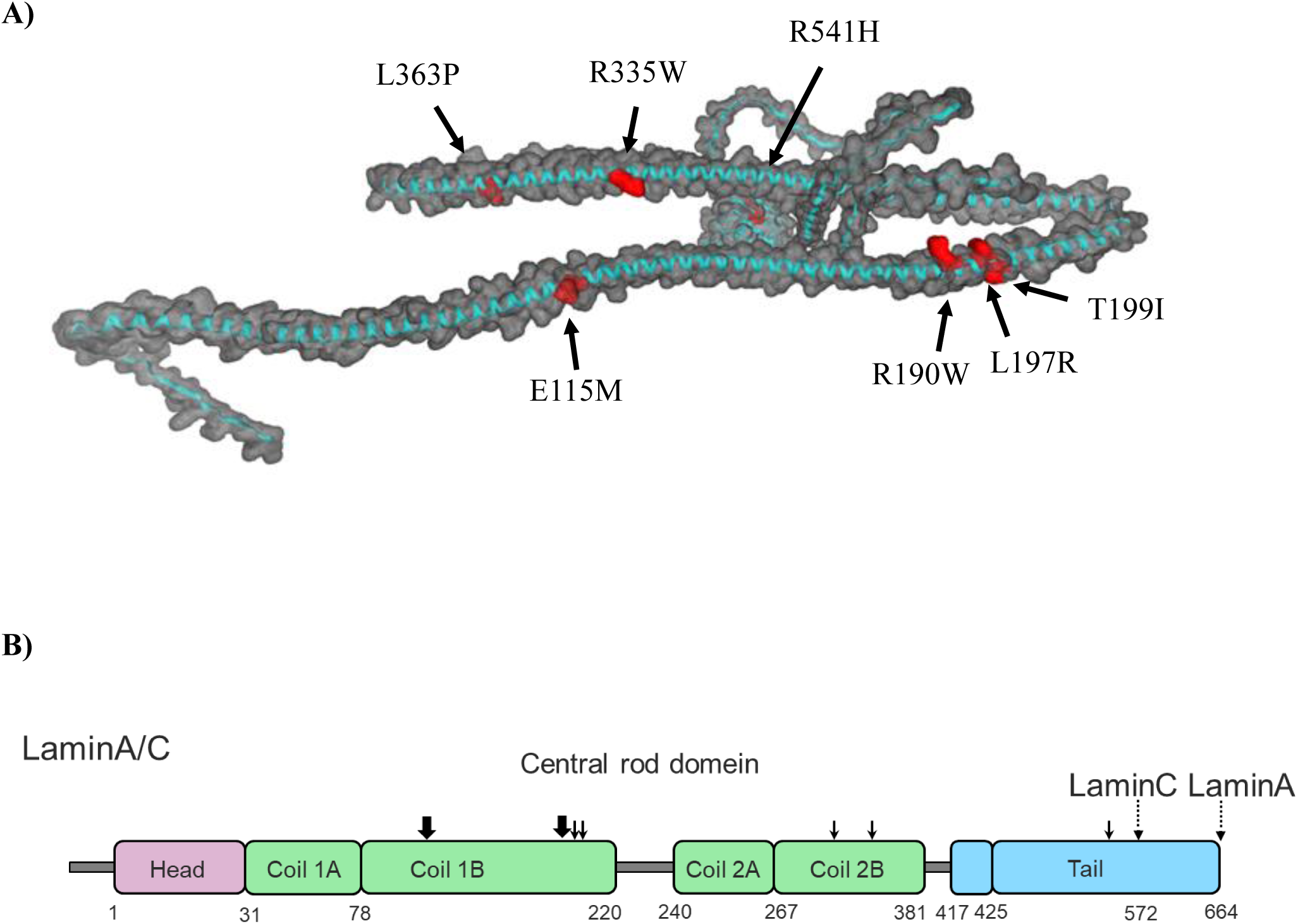
Locations of seven amino acid substitutions in patients with *LMNA* URVs. (A) Location of seven LMNA amino acid substitutions (red) identified in ten DCM patients, mapped onto the 3D structure predicted by AlphaFold2. (B) Schematic of LMNA gene and protein domains. Dotted arrows indicate Lamin A/C regions; solid arrows mark mutation sites (residues 115 and 190 in bold were each found in three patients; residues 197 and 199 in one patient each).

Electron and optical microscopy images of cardiomyocytes were available in nine out of ten patients in (+)LMNA and eight of the ten control cases (**Figure 3**, **Figure 4**, **Figure 5**). Among those in (+)LMNA, electron microscopy images showed only mild structural changes in the nuclei of cardiomyocytes in two cases (**Figure 3H, 3I**), while seven other cases presented pronounced nuclear abnormalities. In the nine cases harboring *LMNA* URVs, cytoplasmic vacuolization was noted in seven; lipid droplets were present in five cases, and lipofuscin deposition in all nine. Additionally, seven cases showed an increased number of mitochondria around the nucleus and in the intermyofibrillar spaces, although no mitochondrial structural abnormalities were detected. In contrast, while mild (**Figure 4D**) or severe (**Figure 4E**) morphological changes in the nuclei of cardiomyocytes were occasionally observed in two control cases without *LMNA* variants, no significant abnormality of the nuclei was observed in the other six cases (**Figure 4**). The optical microscope images of myocardial tissue in (+)LMNA patients showed marked nuclear dysmorphisms in cardiomyocytes but not in other cells, while morphological abnormalities were not observed in the nuclei of cardiomyocytes or other cells in control cases (**Figure 5**).

**Figure 3.**
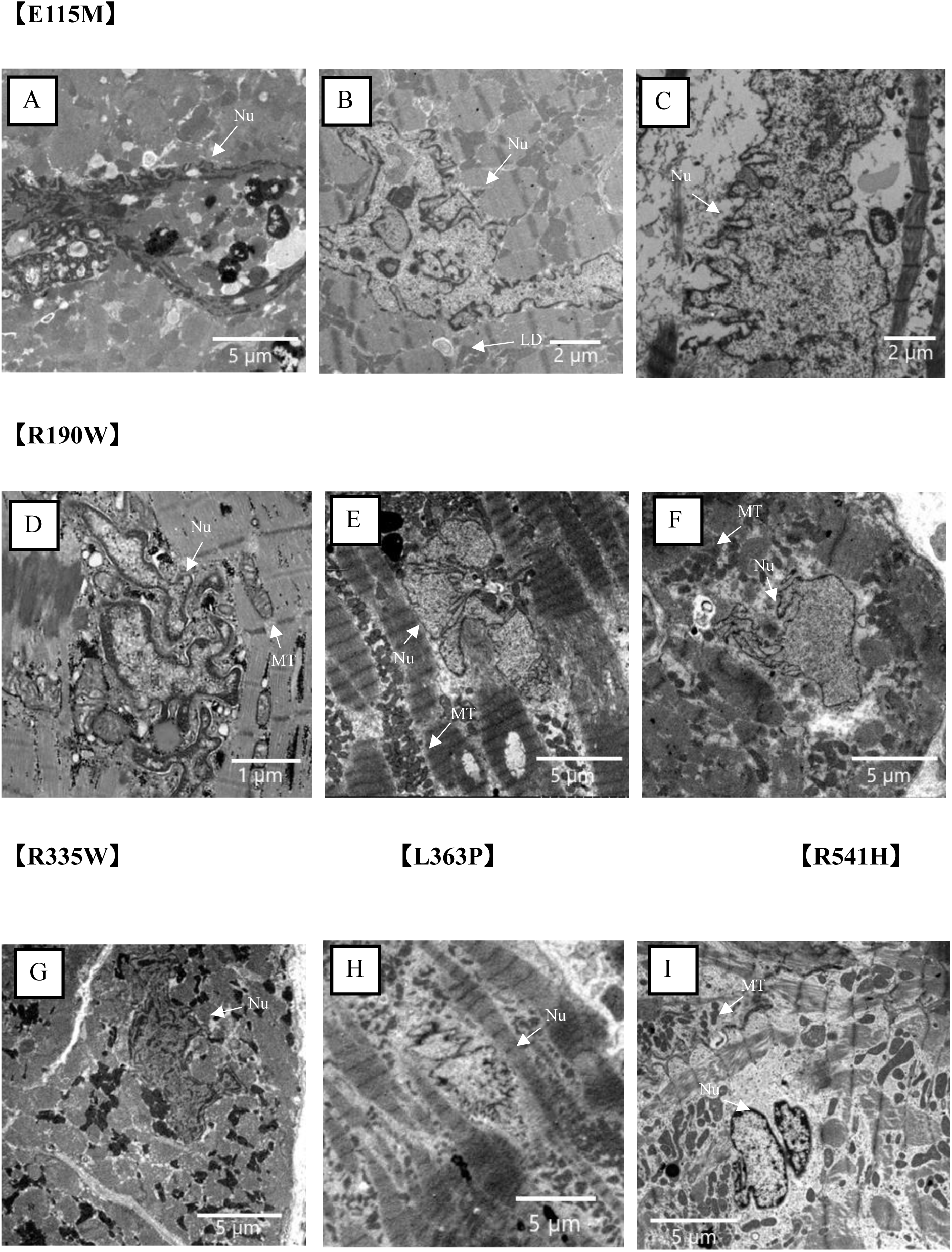
Nuclear morphological abnormalities in cardiomyocytes from patients with *LMNA* URVs. Transmission electron microscopic images of endomyocardial biopsy specimens from DCM patients with *LMNA* URVs. (A–C) E115M; (D–F) R190W; (G) R335W; (H) 363P; (I) R541H. Pronounced nuclear abnormalities of cardiomyocytes are observed in all panels except H and I, which show only mild alterations. Nu, nucleus; Mt, mitochondria; LD, lipid droplets.

**Figure 4.**
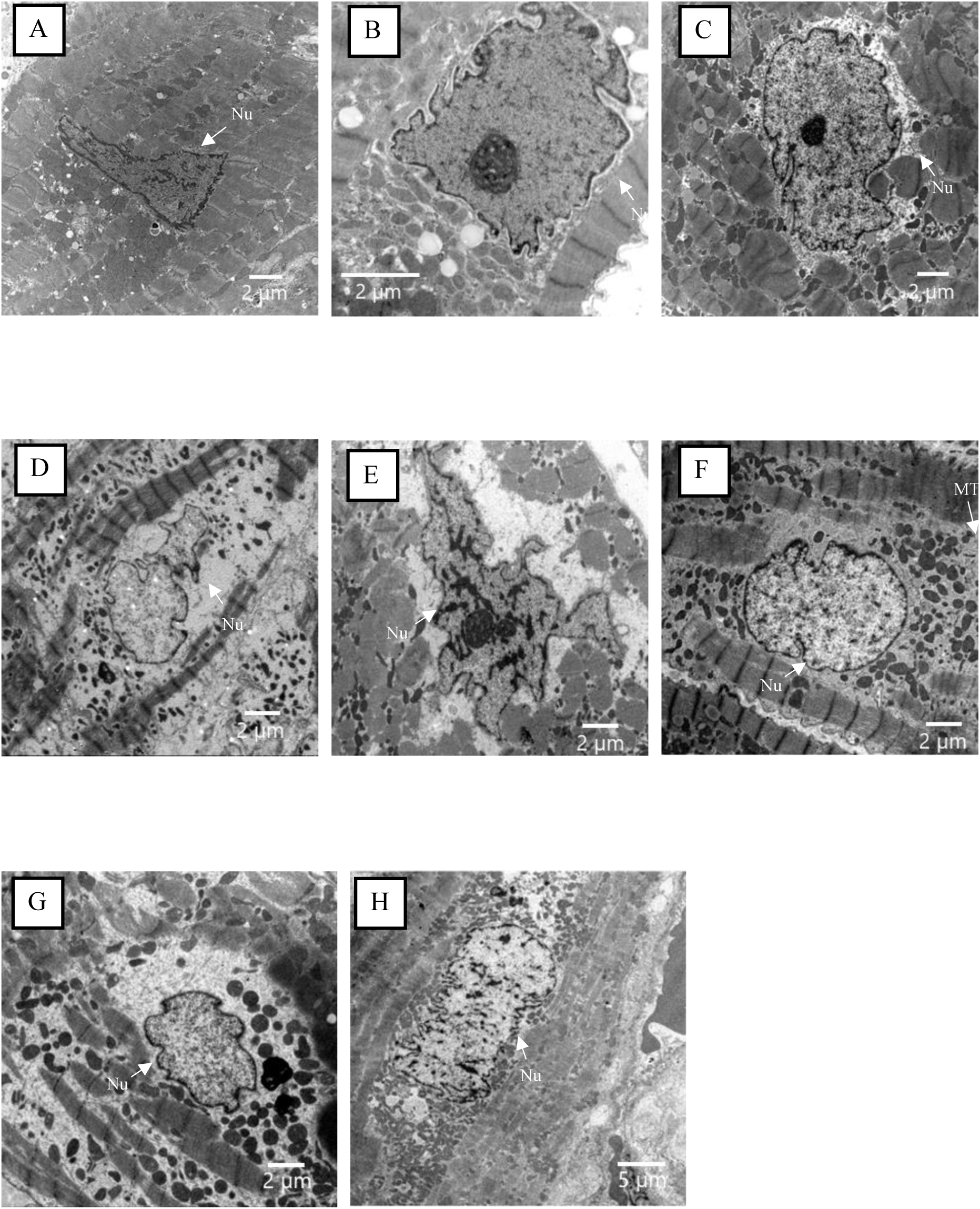
Ultrastructural nuclear morphology in control cases. Transmission electron microscopic images of endomyocardial biopsy specimens from control cases. (A–C, F-H) no significant nuclear changes. (D, E) occasional nuclear morphological changes. Nu, nucleus; Mt, mitochondria; LD, lipid droplets.

**Figure 5.**
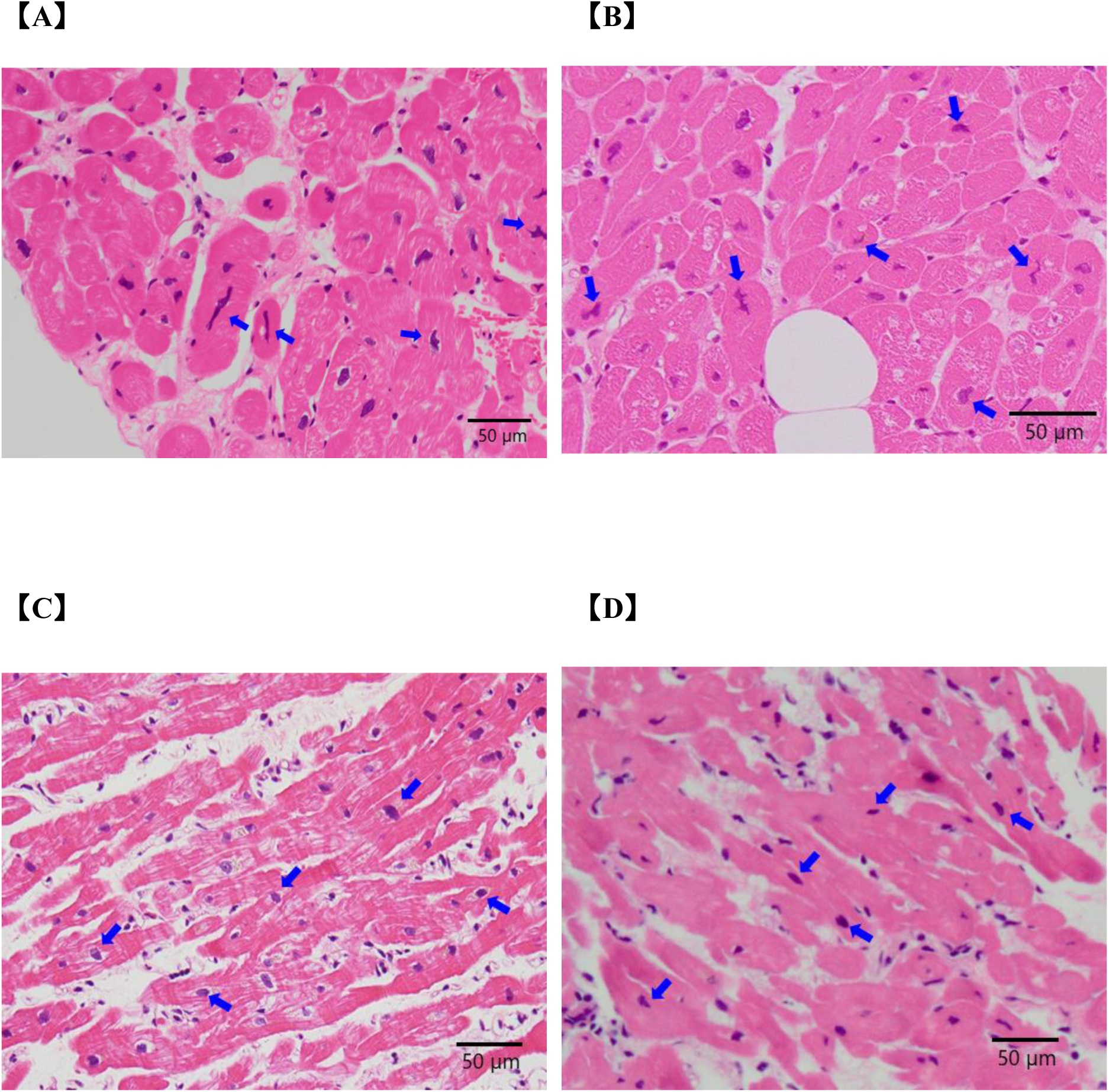
Optical microscopic images of endomyocardial biopsy specimens. Hematoxylin–eosin–stained images of (A, B) DCM patients with *LMNA* URVs (same cases as Fig. 3B and 3F, respectively) exhibiting abnormally shaped nuclei (blue arrows) and (C, D) Control cases (same cases as Fig. 4D and 4F, respectively) showing normal nuclear morphology.

## DISCUSSION

In this study, we systematically investigated the role of coding URVs in DCM utilizing WGS data from 245 patients. Leveraging AlphaMissense pathogenicity prediction and protein-truncating variant analysis, we identified pathogenic coding URVs in 17.6% of patients, consisting of missense variants in *LMNA* and *SLC51A*, and protein-truncating variants in *TTN*. Notably, patients with *LMNA* URVs had worse clinical outcomes and pronounced morphological abnormalities in the nuclei of cardiomyocytes from patients with *LMNA* URVs, suggesting that ultra-rare *LMNA* variants may clinically contribute to disease pathogenesis and could represent important targets for future therapeutic interventions.

To our knowledge, this is among the first studies to link ultra-rare *LMNA* variant with both clinical outcomes and nuclear structural changes in DCM. While several studies have explored the significance of rare variants in various diseases, including cardiovascular disorders,^15,22,23^ URVs have been systematically investigated in only a limited number of conditions, such as schizophrenia.^24^ In the cardiovascular field, URV studies have typically been limited to small-scale analyses, familial studies, or case reports.^25–29^ Although some URV-focused investigations have been conducted in hypertrophic cardiomyopathy,^26^ comprehensive analyses in DCM have remained scarce.^30^ In our study, we identified pathogenic URVs in three genes, *LMNA*, *SLC51A*, and *TTN*, in 43 patients (17.6%) with DCM, a frequency comparable to that reported in previous studies.^27,28^ Among the genes identified in this study, *TTN* and *LMNA* were the most frequently observed, consistent with their known association with DCM.^31^ To explore genotype-phenotype relationships, patients were categorized into three groups: (+)TTN, (+)LMNA, and (-)URVs. Clinical outcomes differed across groups, with *LMNA* URV carriers showing the highest risk of adverse events, including the primary composite endpoint, ICD implantation, and HF hospitalization. These findings align with prior studies showing that *LMNA* variant carriers are at high risk for life-threatening ventricular arrhythmias, regardless of LVEF.^32,33^ Moreover, rare variants in DCM-associated genes have been reported to be more frequently classified as pathogenic in advanced cases requiring LVAD or transplantation than in milder cases.^27^

We investigated *LMNA* to gain insights into the relationship between DCM and genetic rare variants, focusing on nuclear structural changes in myocardial tissue. Optical or electron microscopy images on myocardial specimens were available for examination from nine patients with *LMNA* URVs cases, except for one patient with L197R and T199I substitutions. For comparison, microscopy images from eight control cases without any pathogenic variant in *LMNA* were used. The optical microscopy images revealed significant abnormal morphology of the nuclei in cardiomyocytes from patients with *LMNA* URVs cases, while no such abnormalities were observed in other cell types. Electron microscopy images identified pronounced nuclear abnormalities in seven of the nine *LMNA*-URV cases, whereas mild changes were observed in the remaining two. In contrast, no significant abnormalities were observed in nuclear morphology among six out of eight patients, while mild to moderate changes were observed in two cases in the control cases. These results may represent the first evidence that the *LMNA*-URVs induce morphological abnormalities in the nuclei of cardiomyocytes in patients with DCM.

This study identified seven amino acid substitutions among the ten patients with *LMNA* URVs, six of which were located in coil 1B and 2B of the gene. Considering that the 1B and 2B regions modulate the elasticity of lamin,^34,35^ the *LMNA* URVs in these regions may cause fragility to mechanical stress and disruption of nuclear morphology in contracting tissues, including myocardium. Furthermore, these morphological abnormalities of the nuclei may contribute to impaired nuclear integrity, altered gene expression, and increased cellular stress in cardiomyocytes, ultimately leading to myocardial dysfunction. In short, the *LMNA* URVs may cause fragility of the nuclear membrane, disruption of nuclear envelope morphology under mechanical stress, and finally myocardial dysfunction under the impaired nuclear-cytoskeletal interaction. These findings support a hypothesis that nuclear envelope integrity contributes to cardiac function under mechanical stress.

The selection of allele frequency thresholds for defining pathogenic variants is widely recognized as a critical factor in variant interpretation. In our study, we used a MAF threshold of ≤0.001% for the primary analysis, leading to the identification of three pathogenic URVs. A sensitivity analysis using a more lenient cut-off of ≤0.01% identified additional pathogenic allele counts, but these appeared to have lower pathogenic relevance (**Supplemental Table 2**). Further, at this threshold, only *LMNA* and *TTN* variants were identified as pathogenic mutations, whereas *SLC51A* was not detected. These findings suggest that the cut-off of ≤0.001% purified highly pathogenic variants in this study.

Both E115M and R190W were identified in three patients each in this study, corresponding to a prevalence of 1.3% per variant. Given the ultra-rare nature of these variants in general populations, the identification of each variant in multiple unrelated individuals with DCM strongly supports a pathogenic role. Notably, while R190W is reported as a pathogenic variant associated with DCM regardless of ethnicity, it is registered in public databases such as ClinVar.^36–39^ In contrast, E115M is not listed in such databases and has only been reported in studies specific to Japanese populations.^30^ A previous report identified E115M in three out of 120 DCM cases (2.5%) as a variant of uncertain significance.^30^ In contrast, this study identified that the E115M variant was found in three of 245 patients (1.2%) with DCM as a pathogenic URV based on its high AlphaMissense pathogenicity score, confirmed by the presence of severe morphologic abnormalities in the nuclei of cardiomyocytes and worse clinical outcomes. This identification of a potentially population-specific variant (E115M) highlights the value of ethnicity-tailored genetic analysis and the need for further studies to evaluate its prevalence and pathogenicity across diverse populations.

Of the three genes identified, *LMNA* and *TTN* genes are well-documented in DCM,^40–43^ the roles of *SLC51A* remain insufficiently understood. *SLC51A* has been reported to play roles in the intercellular transport of bile acid and vesicular transport.^44^ Further studies are required to confirm the contribution of these genes to the pathogenesis of DCM. This study identified genes strongly associated with DCM in Japanese patients and highlighted the potential role of *LMNA* variants, including R190W and E115M, in DCM pathogenesis. These findings suggest that *LMNA* variants are significant risk factors in Japanese patients. Early detection of these URVs could serve as a foundation for improving prognosis prediction and treatment strategies for DCM.

### Strengths and Limitations

This is the first study to systematically investigate the contribution of coding URVs to the genetic architecture of DCM using WGS data in Japan. In addition, the use of AlphaMissense, a highly accurate genetic analysis tool, to assess the pathogenicity of URVs brings us closer to understanding the genetic basis of DCM pathogenesis. Our comprehensive approach enabled the unbiased detection of both well-established pathogenic mutations and novel, population-specific variants that are often overlooked by conventional panel-based sequencing. Although our findings are limited to a single ethnic group, the study may address a critical gap in the global literature and generate hypotheses that merit validation in more diverse populations.

The limited number of samples, including controls, used for genetic analysis might have restricted our ability to draw a definitive conclusion, especially concerning the specificity of URVs to DCM. Notably, relaxing the significance thresholds for URVs enabled the detection of variants in genes previously reported to be associated with DCM, suggesting a similar trend as observed in other populations (**Supplemental Table 3**).

Among the three candidate genes analyzed, *SLC51A* genes, excluding *TTN* and *LMNA* harboring URVs, have yet to undergo functional validation, indicating the need for further experimental investigation. Additionally, since no functional assessments of the identified mutations have been performed, further studies are required to determine the pathogenic significance of these variants. Moreover, our pathological findings focused exclusively on URVs in *LMNA*; however, not all DCM patients or cell types were evaluated. It should be noted that the histological images used in this study were obtained in routine clinical practice rather than under a predefined research protocol. Consequently, they may contain incomplete information and be subject to bias, and thus warrant more detailed, protocol-driven analysis in the future. Therefore, comprehensive investigations are warranted to clarify the genes and phenotypes in DCM.

## CONCLUSIONS

Our findings demonstrate the clinical significance of detecting ultra-rare *LMNA* variants in DCM patients. We identified three genes associated with DCM, with *LMNA* variants, particularly E115M and R190W, linked to adverse outcomes and nuclear structural abnormalities. Incorporating URV analysis into clinical workflows could improve risk stratification, enable more accurate prognostication, and facilitate personalized therapeutic interventions, ultimately enhancing outcomes for DCM patients.

## Data Availability

All data referenced in the manuscript is available

## ACKNOWLEDGEMENTS

We thank all the staff of the National Cerebral and Cardiovascular Center, Japan, particularly those in the Department of Heart Failure and Transplantation and the Biobank. We also thank Mie Ekari (Kyoto University) for assistance with AlphaFold2 structure prediction analyses.

The infrastructure of Omics Science Center Secure Information Analysis System, Medical Institute of Bioregulation at Kyushu University provides the (part of) computational resource (https://sis.bioreg.kyushu-u.ac.jp/). This work was supported in part by the MEXT Cooperative Research Project Program, Medical Research Center Initiative for High Depth Omics, and CURE:JPMXP1323015486 for MIB, Kyushu University.

## SOURCE OF FUNDING

This study was supported in part by the Grants-in Aid from the Ministry of Education, Culture, Sports, Science, and Technology to Y.S. (24K02454) and the Agency for Medical Research and Development (AMED), Tokyo, Japan to Y.S. (21ek0210146h9902, 21tm0724602h0001, 22ek0210146h9903, 22tm0724604h0001, 23ek0210194h0001, 24ek0210194h0002, 25ek0210194h0003). M.N. received grants from AMED (JP20ek0109492, JP21wm0425009, JP20ek0109485, JP21ek0109548, JP23ek0109675, JP23ek0109672, JP22tm0424222, JP24gm2010001) and JST NBDC Grant Number JPMJND2302, and JSPS KAKENHI Grant Number JP21H02681. This work was partially supported by the “Joint Usage/Research Center for Interdisciplinary Large-scale Information Infrastructures” and “High Performance Computing Infrastructure” in Japan (Project ID: jh200047-NWH, jh210018-NWH, jh220014, jh230016, jh240015 and jh250010).

## Supplemental Material

**Supplemental Figure 1.**
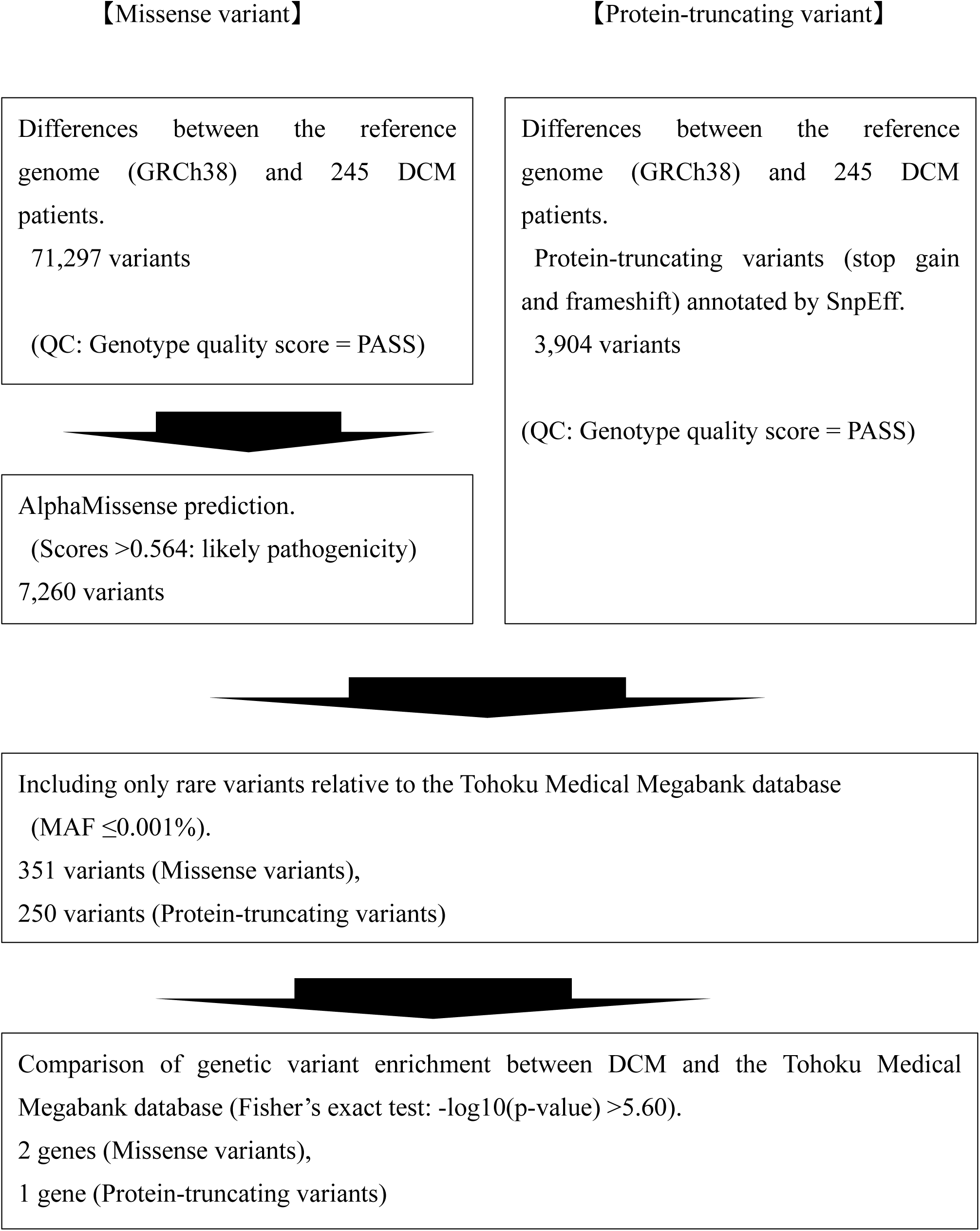
Flowchart for variant filtering and categorization.

**Supplemental Table 1.** Clinical outcomes of *SLC51A* URVs.

|  | *SLC51A* all<br>(N=3) | *SLC51A*<br>(N=1) | *SLC51A* + *LMNA*<br>(N=1) | *SLC51A* + *TTN*<br>(N=1) |
| --- | --- | --- | --- | --- |
| Primary endpoint | 2 | 0 | 1 | 1 |
| - CV death | 1 | 0 | 0 | 1 |
| - HTx | 1 | 0 | 1 | 0 |
| - LVAD implantation | 1 | 0 | 1 | 0 |
| All-cause death | 1 | 0 | 0 | 1 |
| HF hospitalization | 3 | 1 | 1 | 1 |
| ICD or CRT-D implantation | 3 | 1 | 1 | 1 |

Primary endpoint was a composite of cardiovascular death (CV death), heart transplantation (HTx), or left ventricular assist device (LVAD) implantation.
Abbreviations: CV, cardiovascular; HTx, heart transplantation; LVAD, left ventricular assist device; ICD, implantable cardioverter-defibrillator; CRT-D, cardiac resynchronization therapy defibrillator.

**Supplemental Table 2.**
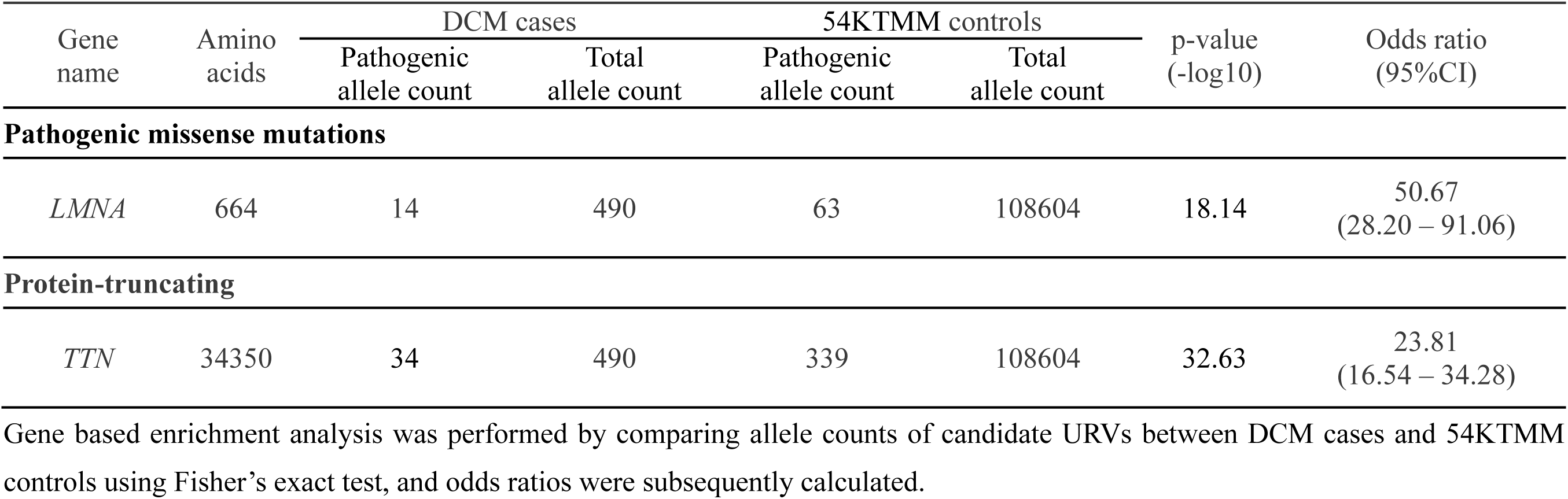
Genes Enriched in DCM Based on URVs with MAF ≤0.01%.

**Supplemental Table 3.** Genes Nominally Associated with DCM Based on URVs with MAF ≤0.001%.

| Gene name | Amino acids | DCM cases |  | 54KTMM controls |  | p-value (-log10) | Odds ratio (95%CI) |
| --- | --- | --- | --- | --- | --- | --- | --- |
|  |  | Pathogenic allele count | Total allele count | Pathogenic allele count | Total allele count |  |  |
| Pathogenic missense mutations |  |  |  |  |  |  |  |
| LMNA | 664 | 14 | 490 | 20 | 108604 | 23.84 | 159.68<br>(80.18 – 318.01) |
| MYH7 | 1935 | 4 | 490 | 46 | 108604 | 4.11 | 19.42<br>(6.97 – 54.17) |
| TNNT2 | 298 | 2 | 490 | 8 | 108604 | 3.05 | 55.63<br>(11.78 – 262.66) |
| ACTN2 | 894 | 2 | 490 | 23 | 108604 | 2.25 | 19.35<br>(4.55– 82.29) |
| Protein-truncating |  |  |  |  |  |  |  |
| TTN | 34350 | 32 | 490 | 163 | 108604 | 39.17 | 46.48<br>(31.47 – 68.65) |
| LMNA | 664 | 2 | 490 | 4 | 108604 | 3.53 | 111.27<br>(20.33 – 608.93) |

Gene based enrichment analysis was performed by comparing allele counts of candidate URVs between DCM cases and 54KTMM controls using Fisher's exact test, and odds ratios were subsequently calculated.

## Notes

### Competing Interest Statement

The authors have declared no competing interest.

### Author Declarations

The study protocols of the NCVC Biobank study and the present study were approved by the ethics committee of the NCVC (Research Approval Number: M23-068 and R21059, respectively).

